# Factors associated with prolonged viral shedding and impact of Lopinavir/Ritonavir treatment in patients with SARS-CoV-2 infection

**DOI:** 10.1101/2020.03.22.20040832

**Authors:** Dan Yan, Xiao-Yan Liu, Ya-nan Zhu, Li Huang, Bi-tang Dan, Guo-jun Zhang, Yong-hua Gao

**Author notes:** Corresponding author: Dr Yong-hua Gao, Department of Respiratory and Critical Care Medicine, The First Affiliated Hospital of Zhengzhou University, Zhengzhou, Henan, China.

## Abstract

**Background:** The duration of viral shedding is central to guide decisions around isolation precautions and antiviral treatment. However, studies about risk factors associated with prolonged SARS-CoV-2 shedding and the potential impact of Lopinavir/Ritonavir (LPV/r) treatment remain scarce.

**Methods:** In this retrospective study, data were collected from all SARS-CoV-2 infected patients who were admitted to isolation wards and had RT-PCR conversion at the NO.3 People’s hospital of Hubei province between 31 January and 09 March 2020. We compared clinical features and SARS-CoV-2 RNA shedding between patients with LPV/r treatment and those without. Logistic regression analysis was employed to evaluate risk factors associated with prolonged viral shedding.

**Results:** Of 120 patients, the median age was 52 years, 54 (45%) were male and 78 (65%) received LPV/r treatment. The median duration of SARS-CoV-2 RNA detection from symptom onset was 23 days (IQR, 18-32 days). Older age (odd ratio [OR] 1.03, 95% confidence interval [CI] 1.00-1.05, p=0.03) and lack of LPV/r treatment (OR 2.42, 95% CI 1.10-5.36, p=0.029) were independent risk factors for prolonged SARS-CoV-2 RNA shedding in multivariate logistic regression analysis. The median duration of viral shedding was shorter in the LPV/r treatment group (n=78) than that in no LPV/r treatment group (n=42) (median, 22 days vs. 28.5 days, p=0.02). Only earlier administration of LPV/r treatment (≤10 days from symptom onset) could shorten the duration of viral shedding.

**Conclusions:** Older age and lack of LPV/r treatment were independently associated with prolonged SARS-CoV-2 RNA shedding in patients with COVID-19. Earlier administration of LPV/r treatment could shorten viral shedding.

**Take home message:** Risk factors for prolonged SARS-CoV-2 shedding included older age and lack of Lopinavir/Ritonavir treatment. Earlier administration of Lopinavir/Ritonavir treatment could shorten the duration of SARS-CoV-2 RNA shedding.

## Introduction

Since first case reported in Wuhan in December, a novel coronavirus, termed SARS-CoV-2, has rapidly spread through other regions of China and the whole world subsequently with 209,839 cases and 8,778 deaths globally as of 19 March 2020 [1-5]. The World Health Organization (WHO) has therefore declared that SARS-CoV-2 has caused a pandemic at 12 March 2020 in view of outbreak of SARS-CoV-2 outside of China. The spectrum of SARS-CoV-2 infection (designated as coronavirus disease 2019 [COVID-19]) ranging from asymptomatic to fatal pneumonia occurred more frequently in patients with underlying conditions [2-7]. To date, there is no vaccine and proven therapy to prevent and treat this deadly virus.

In past two months, there are numerous epidemiological and clinical reports on COVID-19 [2-7]. However, few studies have evaluated the duration of viral shedding which has important implications for guiding decisions around isolation precautions and antiviral treatment in patients with confirmed COVID-19 [7, 8]. Factors associated with prolonged duration of viral shedding are totally unclear. One recent randomized controlled trial showed that Lopinavir/Ritonavir (LPV/r) treatment could not provide incremental benefits, including viral shedding, beyond standard care in hospitalized severely ill patients with COVID-19 [9]. But a post hoc subgroup analysis found that earlier administration of LPV/r treatment could accelerate clinical recovery and reduce mortality [9]. Therefore, it remains crucial to determine whether adding LPV/r treatment impacts the duration of SARS-CoV-2 RNA shedding in non-critically ill patients and whether earlier administration of LPV/r could shorten the duration of viral shedding.

Thus, the study aims to assess the risk factors associated with prolonged viral shedding and the potential impact of earlier administration of LPV/r treatment on the duration of viral shedding in hospitalized non-critically ill patients with SARS-CoV-2 infection during 31 January to 09 March 2020.

## METHODS

### Participants and data collection

This retrospective study included all patients who were laboratory-confirmed SARS-CoV-2 infection and had the available RNA viral data to estimate the duration of viral shedding admitted to the NO.3 People’s Hospital of Hubei province between 31 January 2020 and 9 March 2020, which was one of designated hospital during the outbreak of SARS-CoV-2 in Wuhan. Demographic, clinical, laboratory, treatment and successive viral data were extracted from electronic medical records using a standardized data collection sheet modified based on the WHO/International Severe Acute Respiratory and Emerging Infection Consortium case record form. We assessed severity of illness according to the Chinese management guideline for COVID-19 (version 6.0) [10]. We collected the dose and duration of LPV/r treatment. All data were double checked by two physicians (DY and XYL) with discrepancy resolved by consensus discussion (DY, XYL and YHG). The institutional review board of the NO.3 People’s Hospital of Hubei province approved the study, and patient-level informed consent were waived.

### Virologic investigations

Laboratory identification of SARS-CoV-2 infection was made at Jinyintan Hospital and the NO.3 People’s hospital of Hubei province by real-time reverse transcription-polymerase chain reaction (RT-PCR) assay using the same method described in previous studies with detection reagents provided by the local Center for Disease Control (CDC) [2,3]. After admission, throat-swab specimens were collected and sent to re-detect SARS-CoV-2 RNA by real-time PCR every other day after symptom remission (including fever, cough and dyspnea etc.), and the quantitative viral data was not available. The duration of viral shedding was defined as the date of symptom onset to the day when SARS-CoV-2 was undetectable from two consecutive throat-swab specimens, without a positive test afterward. Corticosteroid treatment was referred to administer a dose equivalent to 25 mg or more of methylprednisolone per day during hospitalization.

### LPV/r treatment

LPV/r (400mg and 100mg, orally, twice daily) was administered to confirmed cases based on the discretion of the physicians at every isolation ward. The treatment duration was 10 days or more with according to the recommendations by Chinese management guideline for COVID-19 (version 6.0) [10]. The exposure of LPV/r was defined as receiving at least one dosage of drug.

### Statistical analysis

Data were presented as mean (standard deviation, SD) or median (interquartile range, IQR) or number (percentage) as appropriate. We employed the Student’s *t* test, ANOVA test, or their corresponding non-parametrical tests for continuous variables, and the *X*^*2*^ test or Fisher exact test for categorized variables for the comparisons of groups as needed. Univariate and adjusted multivariate logistic regression analysis were used to identify risk factors associated with prolonged duration of SARS-CoV-2 RNA shedding. The prolonged viral shedding was defined as the duration of SARS-CoV-2 RNA shedding longer than 23 days. The cut-off was determined a priori based on median duration of viral shedding in our study. Outcomes were defined as initial time of symptom onset to SARS-CoV-2 RNA negativity, which required undetectable RNA from two consecutive throat-swab specimens and without a positive test afterward. Variables used for analysis of prolonged viral shedding included age, sex, smoking, comorbidities (including diabetes, hypertension and cardiac disease), corticosteroid treatment and LPV/R treatment. All statistical analysis was performed using the SPSS version 22.0 software (IBM). A two-tailed P value of <0.05 was regarded as statistically significant.

## RESULTS

### Patient description

From 31 January 2020 to 9 March 2020, 168 confirmed cases admitted to our hospital. As of 9 March 2020, a total of 48 patients (28.6%) were still tested positive for SARS-CoV-2 RNA, of whom 8 patients (4.8%) have died. 120 (71.4%) had resolution of SARS-CoV-2 shedding and were included in the final analysis. All the respiratory specimens tested were from throat-swabs.

Table 1 showed the main characteristics of 120 included patients. Of these patients, the median age was 52 years, 54 patients (45%) were male, 12 (10%) were current smokers and no patients have died. The common comorbidities included hypertension (32 [26.7%]), diabetes (10 [8.3%]) and cardiac disease (7 [5.8%]). 89 (74.2%), 30 (25%) and 1 (0.8%) patients were categorized as general, severe and critical COVID-19. Regarding laboratory results (data available for 119 patients), white blood cell was below the normal range in 27 (22.7%) and above the normal range in 10 (8.4%). 29 (24.3%) had lymphocyte below 0.8×10^9^/liter. The proportion of abnormal platelet count, creatine level and AST were 5.9%, 3.4% and 27.7%, respectively. 54 patients (45%) received systemic corticosteroid treatment and 78 (65%) received LPV/r treatment. The median hospital length of stay was 21 days.

**Table 1.** Characteristics of 120 Hospitalized Patients with SARS-CoV-2 infection in Wuhan.

|  |  |
| --- | --- |
| Characteristic | Value |
| Age, Y | 52 (35-63) |
| Male sex | 54 (45) |
| Current smoker | 12 (10) |
| Comorbidity |  |
| Hypertension | 32 (26.7) |
| Diabetes | 10 (8.3) |
| Cardiac disease<sup>a</sup> | 7 (5.8) |
| Stroke | 3 (2.5) |
| COPD or asthma | 2 (1.6) |
| Chronic renal insufficiency | 1 (0.8) |
| Malignancy | 7 (5.8) |
| **Disease severity** |  |
| General | 89 (74.2) |
| Severe | 30 (25.0) |
| Critical | 1 (0.8) |
| **Laboratory finding on admission\*** |  |
| White blood cell count, $\times 10^9$ cells/L | 5.65 (4.14-7.33) |
| <4 | 27 (22.7) |
| 4-10 | 82 (68.9) |
| >10 | 10 (8.4) |
| Lymphocyte count, $\times 10^9$ lymphocytes/L | 1.14 (0.81-1.55) |
| <0.8 | 29 (24.3) |
| Platelet count, $\times 10^9$ platelets/L | 195 (144-269) |
| <100 | 7 (5.9) |
| Creatinine level, $\mu\text{mol/L}$ | 61 (52-74) |
| >133 | 4 (3.4) |
| AST level, U/L | 31 (24-42.4) |
| >40 | 33 (27.7) |
| **Treatment** |  |
| Corticosteroid therapy | 54 (45.0) |
| Lopinavir/Ritonavir treatment | 78 (65) |
| Antibiotics | 102 (85.0) |
| High-flow nasal canula oxygen therapy | 21 (17.5) |
| No-invasive mechanical ventilation | 2 (1.7) |
| Invasive mechanical ventilation | 1 (0.8) |
| **Outcome** |  |
| Viral shedding duration, d | 23 (18-32) |
| Hospital length of stay, d | 21 (17-26) |

Abbreviations: AST=aspartate aminotransferase; COPD=chronic obstructive pulmonary disease; d=day IQR=interquartile range.
Data presented as n (%) or median (IQR) unless otherwise noted.
\*Data were available for 119 patients
<sup>a</sup>includes congestive heart disease and coronary atherosclerotic heart disease.

### Duration of viral shedding and risk factors

The median duration of SARS-CoV-2 RNA shedding was 23 days (IQR 18-32 days). Only 5 patients (4.2%) had undetectable SARS-CoV-2 RNA within 10 days, 46 (38.3%) tested negative within 20 days, and 85 (70.8%) tested negative within 30 days from symptom onset. Most patients (86.7%) had undetectable SARS-CoV-2 RNA within 37 days after symptom onset, but a small group of 10 patients had detectable SARS-CoV-2 RNA up to 40 days after symptom onset (figure 1). The median duration of SARS-CoV-2 shedding did not differ significantly among groups (general 23 days vs. severe 26 days vs. critical 28 days, p=0.51).

**Figure 1:**
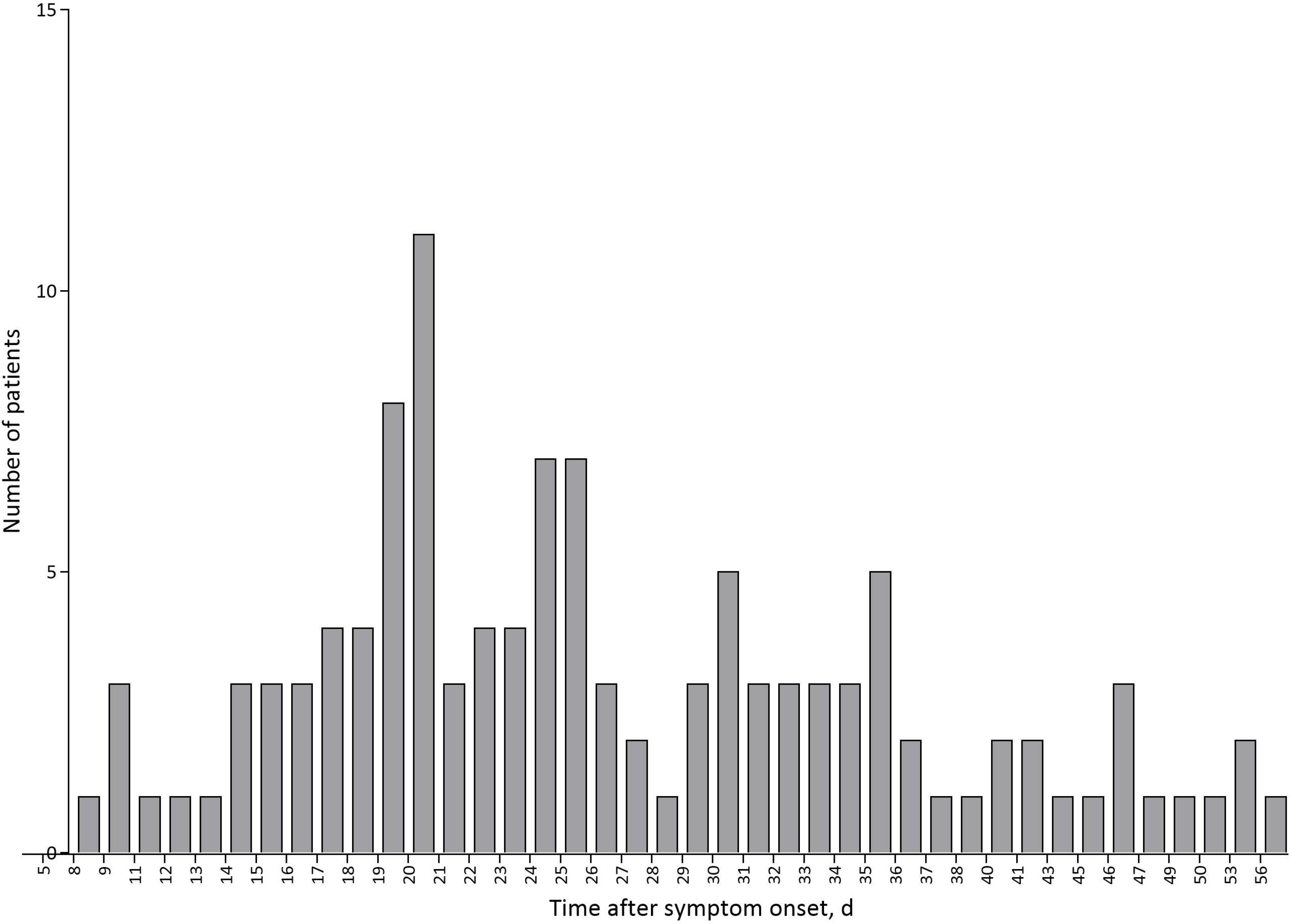
Time to Lopinavir/Ritonavir treatment initiation from symptom onset.

The median duration of SARS-CoV-2 RNA shedding >23 days was defined as prolonged viral shedding in the current study. In the final multivariate logistic model that included all 120 patients, age (OR 1.03, 95% CI 1.00-1.05, p=0.03) and lack of LPV/r treatment (OR 2.42, 95% CI 1.10-5.36, p=0.029) were the independent risk factors associated with prolonged SARS-CoV-2 shedding (Table 2). However, comorbidities (including current smoker, hypertension, diabetes, cardiac disease) and administration of systemic corticosteroid were not associated with prolonged viral shedding.

**Table 2.**
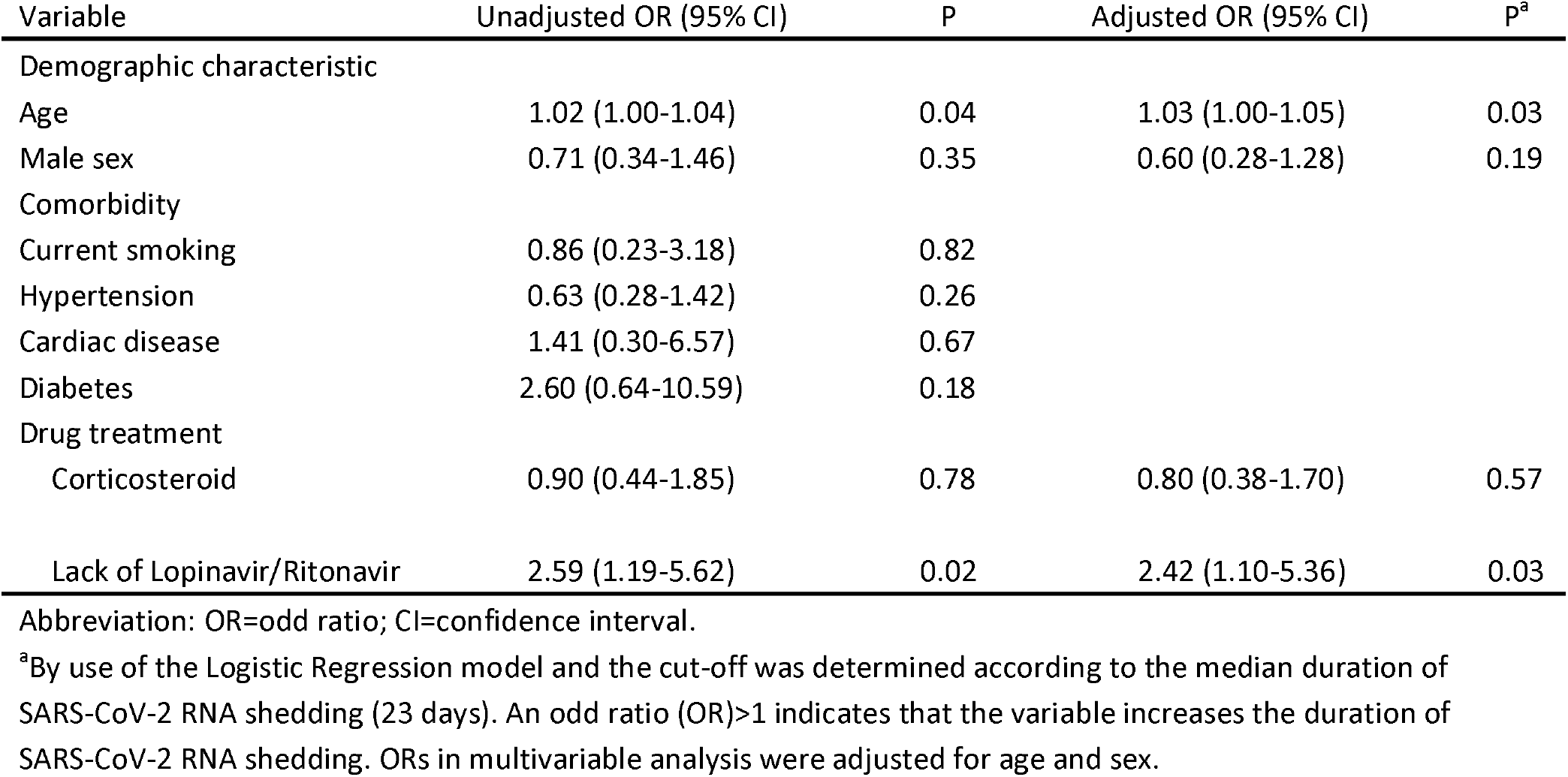
Multivariable Logistic Regression analysis of Factors associated with Duration of SARS-CoV-2 RNA Detection in 120 hospitalized patients in Wuhan

### The effect of Lopinavir/Ritonavir treatment

Of the 120 patients, 78 patients (65%) were administrated with LPV/r treatment. Patients receiving LPV/r treatment and those not receiving LPV/r treatment were similar in most baseline characteristics (Table 3). However, patients receiving LPV/r treatment were more likely to be categorized as severe COVID-19 and have a higher ratio of lymphocyte count <0.8×10^9^/liter than those without LPV/r treatment. LPV/r therapy was started at a median of 10 days (IQR 7-13) from symptom onset. Of 78 patients who received LPV/r treatment, 16 patients (20.5%), 46 (59%) and 64 (82.1%) were initially administered LPV/r treatment within 5 days, 10 days and 15 days from symptom onset, respectively. 7 patients (8.9%) started to administer LPV/r treatment after 20 days (figure 2A). The median duration of LPV/r treatment was 10 days (IQR 9-10). 61 patients (78.2%) received ≥10 days LRV/r treatment. The median duration of SARS-CoV-2 shedding in LPV/r treatment group was 22 days (IQR 18-29), which was shorter than that in no LPV/r treatment group (28.5 days, IQR 19.5-38) (p=0.02). Patients who started LPV/r treatment within 10 days from symptom onset had a shorter duration of SARS-CoV-2 RNA shedding than other patients who began after 10 days (median 19 days vs. 27.5 days, p<0.001). In contrast, the median duration of viral shedding did not differ between patients who initiated LPV/r treatment from symptom onset >10 days and patients who did not receive LPV/r treatment (median 27.5 days vs. 28.5 days, p=0.86) (figure 2B).

**Figure 2:**
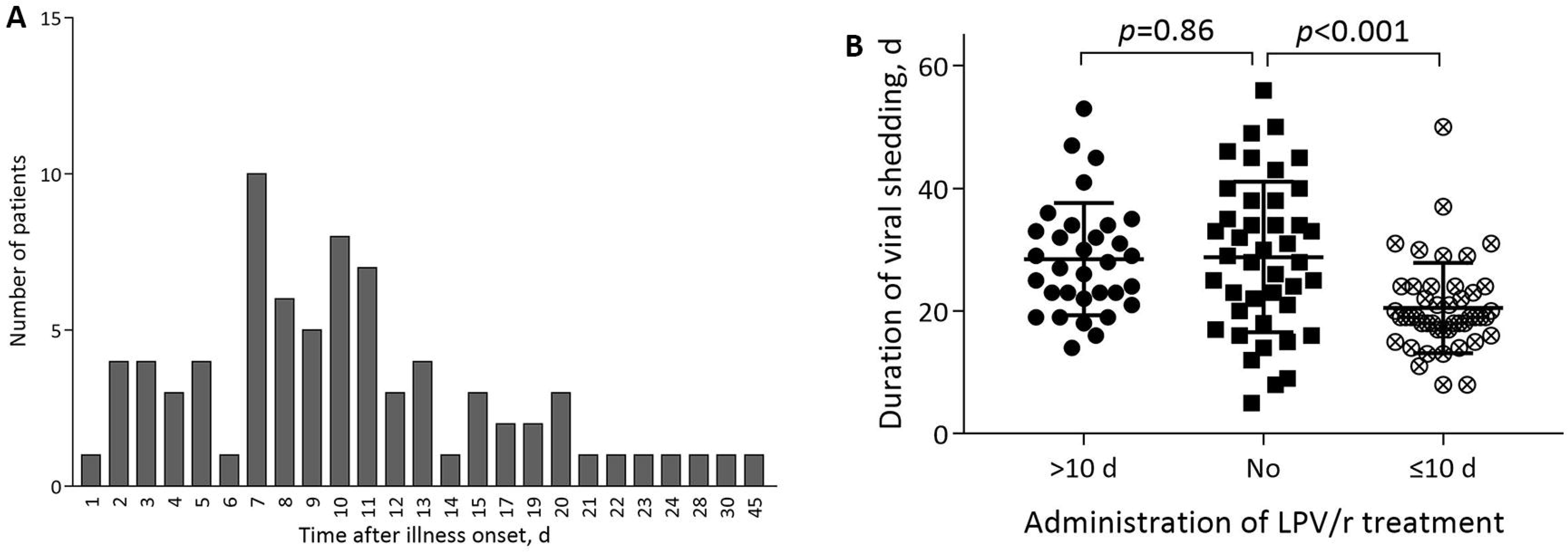
**2A-**Distribution of the number of patients with undetectable SARS-CoV-2 RNA by day after symptom onset; **2B**-Comparisons of the duration of viral shedding among patients who started LPV/r treatment within 10 days from symptom onset, after 10 days, and not received LPV/r treatment.

**Table 3.**
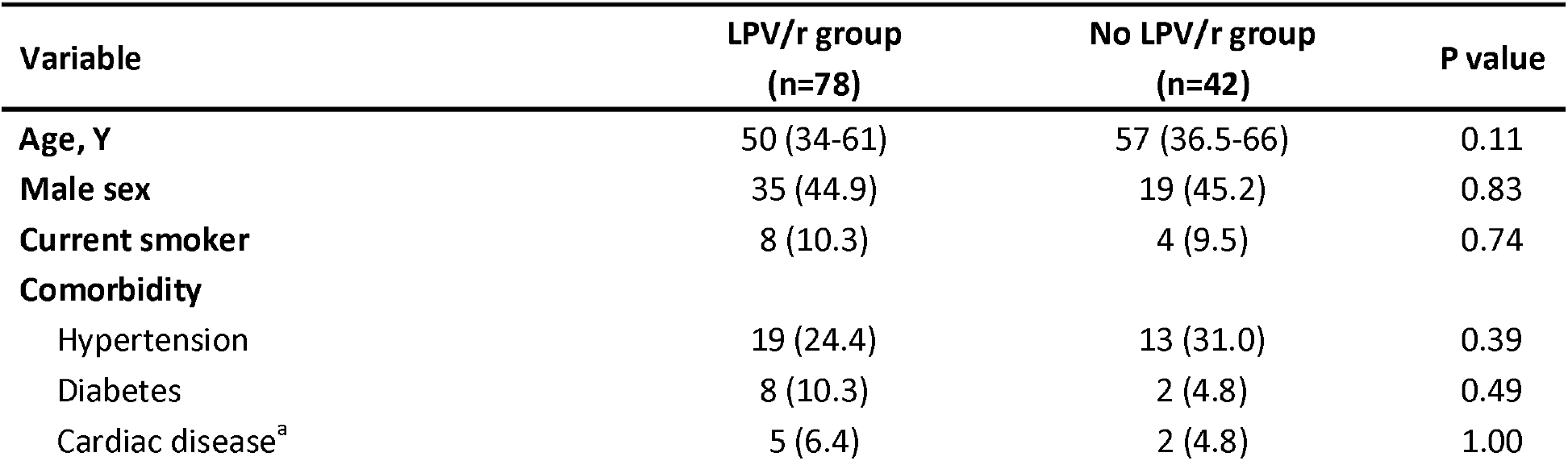
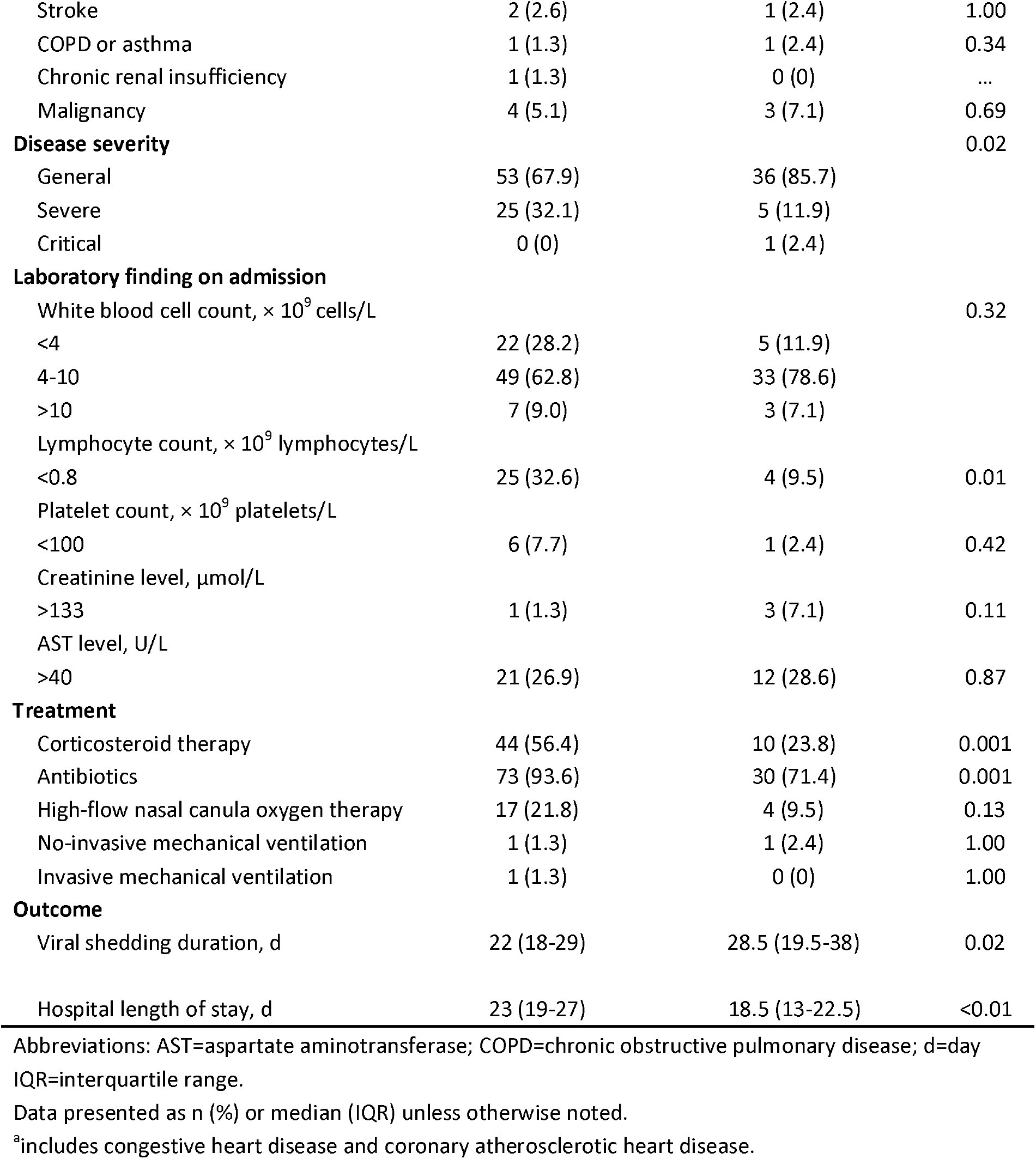
Comparison of clinical features of patients with SARS-CoV-2 infection who treated with Lopinavir/Ritonavir and without Lopinavir/Ritonavir in Wuhan

## Discussion

Viral shedding is commonly used as a proxy measure for infectivity, identification of the duration of viral shedding is central to inform control policies and antiviral treatment in patients with COVID-19. In the current study of 120 hospitalized non-critically ill SARS-CoV-2 infected patients, we, for the first time, identified that older age and lack of LPV/r treatment were the independent risk factors for prolonged viral shedding. We also found that initial administration of LPV/r treatment within 10 days from symptom onset but not afterward could shorten the duration of SARS-CoV-2 RNA shedding. However, we did not observe the impact of comorbidities and corticosteroid use on the duration of viral shedding.

As an emerging novel coronavirus, data on the duration of SARS-CoV-2 RNA shedding are limited. Previously, we reported the median duration of SARS-CoV-2 RNA shedding was 19.5 days [7]. A recent study including 191 confirmed cases showed that the detectable SARS-CoV-2 RNA sustained for a median of 20 days in survivors and persisted until death in non-survivors [8]. Our corresponding estimate in the current study using throat-swab specimen for viral detection was 23 days. About 30% patients had detectable SARS-CoV-2 RNA after 30 days from symptom onset. Investigations of virus shedding in MERS-CoV and SARS-CoV have demonstrated that viral RNA was detected in the respiratory tract specimen for more than 1 month from illness onset [11,12]. Lower respiratory tract (LRT) specimens elicited a higher and more prolonged RNA levels than that in upper respiratory tract (URT) specimens [13]. More severely ill patients typically had higher and more prolonged RNA levels [13]. However, the median duration of SARS-CoV-2 RNA shedding did not differ between severe and general patients in the current study. Previous study reported that detectable SARS-CoV-2 RNA can last until death in non-survivors [8,9]. Whether critically ill patients have more prolonged viral shedding compared to generally or severely ill patients with COVID-19 needs further investigation. To date, studies about the duration of SARS-CoV-2 RNA shedding often used URT specimens for detection [7-9]. Better elucidation of the kinetics of SARS-CoV-2 replication in the upper and lower respiratory tracts in patients with COVID-19 would be useful in future studies.

Our results demonstrated that older age was independently associated with prolonged SARS-CoV-2 RNA shedding. Previous studies have showed that older age is a risk factor for SARS-CoV-2 infection and has been associated with greater risk of development of acute respiratory distress syndrome and mortality [8,14]. Our findings further contributed to the literature that older age was also associated with prolonged viral shedding in patients with COVID-19. One possible explanation is that older patients typically generated less robust innate and adaptive immune limiting viral clearance, and then prolonged the duration of viral shedding [15].

Lopinavir (LPV), a human immunodeficiency virus 1 (HIV-1) protease inhibitor, was combined with ritonavir to increase drug bioavailability of LPV [16]. LPV/r has been showed to be effective in patients infected with SARS-CoV [17]. LPV/r also improved clinical parameters in MERS-CoV infected marmosets and mice [18,19]. Our study, for the first time, showed that lack of LPV/R treatment was independently associated with prolonged SARS-CoV-2 RNA shedding in non-critically ill patients with COVID-19. Administration of LPV/v treatment could shorten the duration of viral shedding compared with no LPV/r treatment. Interestingly, subgroup analysis found that only initial administration of LPV/r treatment within 10 days from symptom onset, but not started after 10 days, could shorten the duration of SARS-CoV-2 RNA shedding compared with no LPV/r treatment. One recent randomized controlled trial showed that LPV/r treatment could not provide additional benefit, including viral shedding, beyond standard care in hospitalized adult patients with severely ill COVID-19 who required a range of ventilatory support mode [8]. However, a post hoc subgroup of the LPV/v in the randomized controlled trial found that earlier administration of antiviral treatment could accelerate clinical recovery and reduce mortality, which is consistent with our findings. Admittedly, patients recruited in the trial were in advanced phase of infection (as evidenced by 25% mortality in the control group). In contrast, most patients (99.2%) in our cohort were non-critically ill patients with COVID-19 due to triage strategies. Because LPV/r is not particularly designed against SARS-CoV-2, it is impossible for LPV/r on SARS-CoV-2 to be as effective as neuraminidase inhibitor on influenza [20]. The median duration of viral shedding even in patients who received LPV/r treatment within 10 days from symptom onset was 19 days in current study, demonstrating that LPV/r treatment could not completely inhibit viral replication. In this regard, earlier administration of LPV/r therapy after symptom onset to patients who were not in late phase of infection may be the key things to determine its efficacy. Nevertheless, randomized clinical trials remain crucial to test whether earlier administration of LPV/r treatment could shorten viral shedding, accelerate clinical recovery and improve survival in patients with COVID-19, especially for non-critically ill patients. One randomized controlled trial for LPV/r in treatment of mild COVID-19 has launched in Korea (NCT04307693).

Our investigation has some limitations. First, owing to triage strategies, almost all patients in our hospital were categorized as general and severe COVID-19, therefore extrapolating these findings to critically ill patients need caution. Second, the estimate duration of SARS-CoV-2 RNA shedding is limited by the type of respiratory specimen, frequency of respiratory specimen collection andlack of quantitative viral RNA detection. Third, the presence of SARS-CoV-2 RNA does not necessarily indicate the production of infectious virus. Forth, we excluded fatal cases from the final analysis because all of them had detectable SARS-CoV-2 RNA until death, and time to death could not accurately reflect the duration of viral shedding, therefore the association between viral shedding and mortality could be not assessed. Finally, interpretation of our findings was limited by sample size. Further large cohort studies are still needed to define the duration of viral shedding and risk factors for prolonged viral shedding in patients with COVID-19.

In summary, older age and lack of LPV/r treatment contributed to prolonged SARS-CoV-2 RNA shedding. Earlier administration of LPV/r treatment can shorten the duration of SARS-CoV-2 RNA shedding. Randomized clinical trials to determine the effectiveness of LPV/r treatment in non-critically ill patients with COVID-19 are still needed.

## Data Availability

We declare that all data are available

## Author contribution

Y.H.G. designed the project. D.Y., X.Y.L and B.T.D. carried out the data collection. Y.H.G., Y.N.Z. and L.H. analyzed the data and prepared the figures. Y.H.G. D.Y. Y.N.Z. and L.H. drafted the manuscript. All the authors have revised the manuscript critically, approved the version submitted for publication and have agreed to be accountable for all aspects of the work.

